# People underestimate the change of airborne Corona virus exposure when changing distance to an infected person: On interpersonal distance, exposure time, face masks and perceived virus exposure

**DOI:** 10.1101/2022.03.14.22272341

**Authors:** Ola Svenson

## Abstract

Participants judged airborne Corona virus exposure following a change of inter-personal distance and time of a conversation with an infected person with and without a face mask. About 75% of the participants underestimated how much virus exposure changes when the distance to an infected person changed. The smallest average face to face distance from an infected person without a mask that a participant judged as sufficiently safe was about 12 feet (3.67 m). Correlations showed that the more a person underestimated the effects of change of distance on exposure the shorter was that person’s own safety distance. On average the effects of different lengths of a conversation on exposure were correct, but those who judged the effects of time as smaller tended to select longer safety distances. Worry of own COVID-19 infection correlated with protective behaviors: keeping longer safety distances, avoiding public gatherings, postponement of meetings with friends. The results showed that the protective effects of both distancing and wearing a face mask were under-estimated by a majority of the participants. Implications of these results were discussed last.

## Introduction

The COVID-19 pandemic caused by the SARS-CoV2 virus is likely to be followed by other pandemics also caused by airborne Corona or other viruses. During the COVID-19 pandemic people were asked to keep distance, wash their hands, avoid gatherings of people etc. But, do people know about the degree to which these measures protect them from virus exposure and infection? Do they know how a shorter distance between two people increases virus exposure? Do people know to what extent the length of a social contact influences virus exposure? Do they know how much a face mask protects them from virus exposure? The present study was focused on how people perceive changes in virus exposure from an infected person when interpersonal face to face distance is changed and when the time for a face to face conversation is changed.

The Covid-19 pandemic illustrates the need of expert communication about risks to the public and policy makers for informed decision making. Based on the information available to them, lay people develop their own mental models about causal and other relationships between different variables. Therefore, we should find out about these models and modify them if they are wrong or represent biased scientific facts. In the present context we will focus on virus exposure in face to face conversations at different distances and lengths with and without a mask. In a conversation between two people one of whom is infected with an airborne virus and the other not, there is an exchange not only of words but also of airborne particles including viruses. In the case of face to face situation, a person needs to know what a deviation from a chosen or a recommended distance means for virus exposure. Only with such knowledge can she or he make informed decisions for informed and persistent protective behaviors.

With a focus on inter individual distances, research questions like those raised above become relevant. To specify, do people know how actual exposure to an airborne virus varies with variation of face to face distance and length of a conversation? What does it mean to a person’s virus exposure to move closer from, e.g., 6 feet to 2 feet from a person who is infected with an airborne virus like the SARS-CoV2? What does it mean to extend a conversation from 1 minute to 5 minutes and how can a face mask change virus exposure? To answer these questions, we asked participants to judge virus exposure after changes of interpersonal distance, length of a conversation and with or without a face mask. The judgments were related to empirical and theoretical facts concerning the spread of virus particles in the air. Judged exposure was also related to perceived risk and some individual characteristics. To exemplify, what individual factors correlate with the accuracy of the participants’ exposure judgments and with their preferences for inter person distance during a conversation?

To the best of our knowledge, there are no studies of perceived exposure as a function of changes of inter person distance except the study by Svenson and colleagues (Svenson et al., 2020) who found that most participants underestimated the protective effect of moving further away from another person. Correspondingly, most participants were unaware of how much their exposure would increase if they moved closer to the other infected person. The present study departed from this study and extended the scope to include time of a conversation with and without a mask personal characteristics and risk perception.

### Objective measures of virus exposure

The distance that particles travel away from an infected person depends on many different factors, such as the size of the particles, initial momentum with which they are expelled (regular conversation, singing coughing etc), position of the head and the body of the person emitting the particles, strength (velocity), structure (turbulent or laminar), direction, temperature and humidity of airflow and individual differences between people. A SARS-CoV2 virus can travel on droplets that are greater or smaller than 5 μm. Droplets that are exhaled from a person and are greater than 5 μm follow the laws of gravity and fall to the ground within some distance from the exhaling person (Morawska & Cao, 2020). Droplets smaller than 5 μm, called aerosols, can originate directly from an exhalation or from evaporated greater droplets and their movements follow the streams of air and can stay in the air for a long time (Crema, 2020; Setti et al., 2020; Lonergan, 2020). The aerosols can provide ambient virus exposure. Early after an exhalation, the ambient dispersion effects can be ignored because the droplet-laden effects seem to dominate, but after some time aerosols accumulate and the ambient effect takes over and determines the exposure if a space is no properly ventilated (Walker et al., 2021). Hence, a person’s exposure to droplets and aerosols depend not only on distance but also on other factors and situations. The present project focused on distance and time of a conversation keeping other variables constant assuming a location for a conversation in an open space with calm air as in a spatious mall or outside with no wind.

A physical theory of virus exposure that can be related to perceived exposure should be based on virus distribution in the air under different conditions. Balachander and co-authors (Balachander et al., 2020) gave an extensive overview and possible solutions for how to solve the multiphase flow problems created by droplets and aerosols carrying viruses. To illustrate, Bourouiba (2020) specified how far the larger droplets but also smaller aerosols can travel after a sneeze or cough (7–8 m). In the present studies we treated only normal breathing conditions. To illustrate further, Bjørn and Nielsen (2002, p. 155, Fig. 15) reported exposure to a another person’s normal breathing in a calm laboratory face to face setting with different distances (0.4 to 1.2 m). The power function Exposure = 1.90 × Emission × Distance ^−2.2^ describes their results. In another empirical study by Nielsen et al. (2012, p. 557, Fig. 8) the power function was Exposure = 4.3 × Emission × Distance ^−2.3^ (0.35 to 1.10 m) with a different constant depending on different measures of relative exposure used in the different studies.

Recently, Melikov (2020) presented an overview of studies with exposure as a function of distance reported by different authors (Ai et al., 2019; Bolashikov et al., 2012; Liu et al., 2016; Olmedo et al, 2012: Olmedo et al., 2013; Villafruela et al., 2016). The results were summarized by Figure 1 in Melikov (2020 p. 2) with a decreasing function that can be approximated by a power function with an exponent that is - 2 or smaller. This was also confirmed by Wang, Xu and Huang (2020). Howard and collaborators (Howard et al., 2021) reviewed research about face masks and protection against droplets and aerosols who can carry corona viruses. In general, they found that high quality face masks filter at least about 90% of droplets and aerosols exhaled by a person.

The laboratory results presented by Nielsen and collaborators and most of the above cited researchers were obtained in conditions similar to the situation presented to the participants in the present and in the study by Svenson and colleagues (Svenson et al., 2020), a face to face conversation with no coughing or sneezing, but a change of inter-personal distance. Therefore, we used the power function in Equation (1) when we formulated a model for objective exposure. We set the exponent to 2.0 in the model. It is clear from the empirical studies cited above that this exponent, if anything, underestimates the speed of decrease of exposure with increasing distance. This allows some margin on the conservative side when comparing subjective judgments with physical facts. In Equation (1), *Epv* is exposure to virus, *a* and *n* are constants, *E* emitted virus during time *t*, and D distance to source.

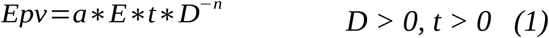

We will call the function in Equation (1) the Virus Exposure Model, VEM. The exponent describes change of exposure as as a function of change of distance. Applying n = 2.0 to a person who approaches another infected person from 6 feet to 2 feet, it predicts an increase of exposure to (6/2)^2^ = 9 times. However, a person who applies a linear model, n = 1.0 will judge the exposure to be 6/2 = 3 times the initial exposure after the approach. The study by Svenson and colleagues (Svenson et al., 2020) on perceived exposure showed that different judgments models and exponents were used across participants. Some participants used a linear model and a majority of the participants underestimated the effects on exposure following both approach and withdrawal from another person.

### Behavior

There are many studies of behavior in general and protective behavior in epidemic or pandemic situations. Some of them study which psychological characteristics and processes correlate with protective behavior on a general level. In a national survey from March 2020, Bruine de Bruin and Bennet (2020) found that perceived COVID-19 risk of infection was significantly and positively correlated with the protective behaviors “washing hands”, “avoiding public spaces”, “avoiding high-risk individuals” and “canceling travels”, but not with the perceived risk of dying from the disease. To exemplify further, Vacondio and colleagues (2021) investigated self-protective behavior in the UK, Italy and Austria. In their study from April 2020, they found that self-reported protective behavior was positively correlated with worry and to some extent mediated by perceived threat from the disease. Shilo, Peleg and Nudelman (2021) studied cognitive and emotional factors and their relationships with reported adherence with behavioral guidelines. The results showed how both emotional factors,and cognitive factors correlated positively with self reported protective behavior. The cognitive factor “efficacy of behavior” was the most prominent factor with R= 0.70, and different kinds of emotional worry had correlation with adherence to behavior guidelines around R= 0.20. The fact that the belief in efficacy of protective behavior in general leads to the question of what components of behavior are perceived to contribute to efficacy including keeping distances. With reference to these results, we included questions about self protective behavior, perceived threat from catching Covid-19, worry and efficacy of protective behavior.

In a judgment study, Lisi and colleagues (Lisi et al., 2021) reported that their participants preferred significantly shorter face to face distances to another person with a face mask compared to a person without a face mask and longer distances if the other person was diagnosed with COVID-19. Other studies show that mask wearing is positively correlated with other preventive measures, including hand hygiene and physical distancing (Howard et al., 2021). Welsch and colleagues (Wesch et al., 2021) asked their participants to indicate the inter-personal distance that they preferred without the Corona virus and during the pandemic and the distances were 1.18 m and 1.83 m. However, the latter distance decreased over time during the pandemic to 1.41 m in some kind of adaptive process. Hall (1966) suggested different interpersonal distances: close distances for partner/ family (up to 0.45 meters), distance to friends (0.45-1.20 meters) and distance to strangers (1.20 -3.65 meters). With reference to these distances and recommended inter-personal distance during the pandemic, we used distances from 2 feet (0.60 m) to 6 feet (1.80 m) in the present study and asked about preferred inter-personal distances with and without a mask on an infected person.

In the first study, participants judged changes in exposure following both inter-personal distance and changes of time of a conversation. Based on earlier results, we predicted that the effects on virus exposure from changing inter-personal distance will be underestimated for both approaching and moving away conditions. Based on the fact that a linear functions is the most easily available tested in a hierarchy of subjective functions we predicted that a majority of the time judgments will be described by a linear function, the first function to be elicited in an unknown situation (Svenson, 2016). Furthermore, we predicted that a person’s shortest acceptable face to face distance to an infected person will co-vary with judgments of exposure following change of distance and time. We predicted that a person who accepts only a longer distance from another infected person to be safe will also judge a change in interpersonal distance to have a smaller effect than a person who accepts a shorter safety distance. This is why the former needs to move further away. We had no well grounded hypotheses about the effects on judged exposure of time of a conversation and face mask investigated in study 2. However, the fact that a linear function is easy to apply and elicit (Svenson, 2016) indicates that judgments of the effect of exposure time will not be systematically biased. Research has shown that worry is one of the important drivers of protective behaviors (Bruine de Bruin & Bennett, 2020; Frounfelker et al.,2021; Shilo, Peleg & Nudelman, 2021; Vacondio et al., 2021) and therefore we predicted that worry should correlate positively with protective behavior also in the present study.

## Study 1: Effects of distance and time on exposure

### Method

#### Participants

In all, 101 participants aged 18 år more were recruited by Prolific from a general US adult English speaking population. One participant was eliminated because of failure on an attention test question and 4 participants were excluded because they did not fulfill the task by giving only zero or 100 as answer to the questions. Hence, the study included 96 participants (48 women, 47 men and 1 unspecified). The mean age was 31 (SD= 11.4) with a range from 19 to 73 years. In the sample 4 had high school no graduate education, 16 high school graduate, 26 some college, 39 college graduate and 11 more than college graduate education.

#### Procedure and problems

A Qualatrics questionnaire was distributed to the participants and on average a participant used 14 min to complete the task. The instruction started with the following.

***“****As you probably know, the Coronavirus spreads on small droplets in the air when a person infected with Covid-19 breaths, coughs, sneezes or talks without wearing a mask. Therefore, keeping a physical social distance reduces any virus exposure and the risk of the virus to spread from person to person. We will ask you to judge the degree to which different distances in face to face situations can reduce exposure to the virus for persons who do not wear a mask*.*”*

The instruction continued with an example that introduced the problems about inter-personal distance. The instruction to the condition with exposure from a longer distance compared with a shorter distances included the following.

*“Assume that two persons are in a face to face conversation in* ***6 minutes standing 2 feet*** *away from each other and one of them is infected by a Corona virus. If they had been* ***further away*** *from each other, for example*, ***6 feet*** *the virus exposure* ***would have been smaller***.

*What percentage of the airborne viruses reaching a person at 2 feet will reach a person at 6 feet? Please, answer with a percentage. Same = 100%, Three quarters = 75%, Half = 50%, One quarter = 25 %, One tenth = 10% etc…. “*

Problems varying the time of a conversation were introduced as follows in the from long to short time condition.

*“Assume that a conversation was 6 minutes and* ***became shorter***. *Then the virus* ***exposure would become smaller***.

*What percentage of the airborne viruses reaching a person during a shorter conversation of* ***2 minutes*** *at* ***3 feet*** *will reach a person compared with the exposure* ***during 6 minutes****? Please, answer with a percentage. Same = 100%, Three quarters = 75%, Half = 50%, One quarter = 25 %, One tenth = 10% etc…. “*

The problems were presented in blocks of 6 problems in each of the conditions: (a) from long to short distance (time constant), (b) short to long distance (time constant), (c) from short to long conversation time (distance constant) and (d) from long to short conversation time (distance constant). All blocks were presented to each participant with either distance or time presented first for half of the participants in a balanced design. Blocks with increasing distance or time were presented first for half of the participants in a balanced order. Hence, the design was 2 x2 (distance/time presentation first x increasing/decreasing presentation first). The items in a block were presented in a unique random order to each participant. The distances and times in the problems are listed in Tables 1 and 2. Some general demographic questions and questions about a participant’s worry over COVID, preferred interpersonal distance and some other questions related to a participant’s experience with COVID followed. Last, we presented three cognitive problems measuring cognitive ability. However, they had no relationship with any of the other items or scales and were not treated further. A complete set of questions can be found in Appendix 1.

**Table 1.** Medians and quartiles of exposure judgments and predictions according to VEM, n= 2.0 and linear model with n= 1.0. Increasing and decreasing distance.

| **In-creasing distance** | **First distance (feet)** | **Second distance (feet)** | **Mean (SD)** | **Judged median exposure % (quartiles)** | **N** | **Predicted VEM n = 2.0** | **Predicted linear n = 1.0** | **Median - VEM predict n = 2.0** | **Judgment - linear predict n = 1.0** |
| --- | --- | --- | --- | --- | --- | --- | --- | --- | --- |
| **1** | 4 | 5 | 62.31<br>(30.16) | 75.0<br>(47.5 - 80.0) | 96 | 64.0 | 80.0 | 11 | -5.0 |
| **2** | 2 | 6 | 32.86<br>(21.52) | 30.0<br>(15.0 - 40.0) | 95 | 11.0 | 33.0 | 19 | -3.0 |
| **3** | 4 | 6 | 55.21<br>(21.48) | 60.0<br>(40.0 - 75.0) | 96 | 44.4 | 66.0 | 16 | - 6.0 |
| **4** | 2 | 5 | 38.25<br>(17.90) | 40.0<br>(25.0 - 50.0) | 95 | 16.0 | 40.0 | 24 | 0.0 |
| **5** | 5 | 6 | 64.60<br>(32.44) | 80.0<br>(30.0 - 98.5) | 96 | 69.4 | 83.3 | 11 | -3.3 |
| **6** | 2 | 4 | 49.92<br>(14.98) | 50<br>(50.0 - 50.0) | 96 | 25.0 | 50.0 | 25 | 0.0 |
| **De-creasing distance** |  |  |  |  |  |  |  |  |  |
| **1** | 5 | 4 | -* | 125<br>(100 - 175) | 85 | 156 | 125 | - 31 | 0.0 |
| **2** | 6 | 2 | - | 300<br>(300 - 500) | 84 | 900 | 300 | - 600 | 0.0 |
| **3** | 6 | 4 | - | 150<br>(150 - 200) | 85 | 225 | 150 | -75 | 0.0 |
| **4** | 5 | 2 | - | 250<br>(215 - 500) | 84 | 625 | 250 | - 375 | 0.0 |
| **5** | 6 | 5 | - | 120<br>(100 - 150) | 84 | 144 | 120 | - 24 | 0.0 |
| **6** | 4 | 2 | - | 200<br>(200 - 313) | 84 | 400 | 200 | -200 | 0.0 |

\* the distributions had too many outliers for reliable means.

**Table 2.** Medians and quartiles of judgments and predictions according to linear model. Decreasing and increasing time of talk.

| **De-creasing time** | **First time (minute s)** | **Second time (minutes)** | **Judged exposure** | **N** | **Predicted linear** |
| --- | --- | --- | --- | --- | --- |
| **2** | 10 | 1 | 10.0<br>(10 - 10) | 96 | 10.0 |
| **3** | 6 | 1 | 17.0<br>(15 - 25) | 96 | 17.0 |
| **4** | 10 | 3 | 30.0<br>(30.0 - 35.0) | 96 | 30.0 |
| **5** | 3 | 1 | 33.0<br>(25.0 - 33.0) | 96 | 33.0 |
| **6** | 10 | 6 | 60.0<br>(50.0 - 61.3) | 96 | 60.0 |
| **In-creasing time** |  |  |  |  |  |
| **1** | 3 | 6 | 200<br>(200 - 200) | 86 | 200 |
| **2** | 1 | 10 | 1000<br>(1000 - 1000) | 90 | 1000 |
| **3** | 1 | 6 | 600<br>(500 - 600) | 88 | 600 |
| **4** | 3 | 10 | 333<br>(300 - 500) | 89 | 333 |
| **5** | 1 | 3 | 300<br>(200 - 300) | 86 | 300 |
| **6** | 6 | 10 | 180<br>(165 - 400) | 85 | 167 |

## Results

Ten of the participants had been infected with the Corona virus. We will include also these participants in the group data analyses because the sample was too small for separate analyses. A number of participants who were asked to judge percentages greater than 100 in a decreasing distance condition (increasing exposure) gave judgments that were smaller than 100. In spite of a detailed instruction, they may have misunderstood the task so that they judged the increment in exposure instead of the total exposure after a change. When a judgment was smaller than 100 in an increasing distance condition (decreasing exposure) the judgment was coded as missing. There were only a few judgments above 100 in the increasing distance (decreasing exposure) conditions and they were also coded as missing. The judgment distributions were all skewed with high skewness (less than - 1.0 or greater than + 1.0) or moderate skewness (between +/- 1.0 and +/- 0.5). Therefore, we focused on medians in the data analyses and excluded means for some conditions in Table 1, because there were too many outliers distorting the means.

### The bias

Table 1 gives medians and quartiles for each of the distance problems. In comparison with the VEM model with n = 2.0, the average participant overestimated exposure for increasing distance and underestimated exposure for decreasing distance. This means that they were not sufficiently sensitive to the effects on exposure of changing distance to an infected face to face speaker.

For both increasing and decreasing distance separately, the differences between the median judgments and predictions of the VEM 2.0 model were significant in Wilcoxon Signed-Ranks tests, T=0, α=0.05, N=6 for both conditions. The numbers of observations were too small for a test based on normal distributions and therefore the z values are not reported. It is clear that the median judgments in Table 1 are closer to a linear function than to VEM 2.0.

We computed the percentages of participants who realized the degree to which exposure changes with changes in distance following the VEM 2.0 function or faster. For the increasing distance problems meaning a decrease in radiation, these percentages were for change from 4 to 5 feet, 40%, 2 to 6 feet, 22%, 4 to 6 feet, 27%, 2 to 5 feet 8%, 5 to 6 feet 33% and 2 to 4 feet 10% with mean 23%. Hence, 77% of the judgments indicated that the participants did not realize how fast virus exposure decreases with increasing distance to an infected person.

For the decreasing distance (increasing exposure) problems the participants who realized how fast exposure increases compared to VEM 2.0 following an approach were from 5 to 4 feet, 28%, 6 to 2 feet, 16%, 6 to 4 feet, 21%, 5 to 2 feet, 11%, 6 to 5 feet, 31% and 4 to 2 feet, 19%.with mean 21%. Hence, 79% of the judgments indicated insensitivity to the fast increase of exposure following an approach towards an infected person.

Next, we investigated the effects on exposure following changes of the length of a conversation. Table 2 shows median judgments for decreasing and increasing time. The results show an almost perfect fit to a linear relation. This means that the participants were quite accurately sensitive to the effects of changing the time of a conversation according to VEM with a constant distance.

In summary, the results replicated the 2020 findings (Svenson et al., 2020) with significant underestimations of the effects on exposure of changing distance. Second, the participants followed an adequate linear model when they judged the effects of time, quite in line with distance constant in Equation (1).

We computed Cronbach’s alfa for each of the four conditions. The six problems in the increasing distance condition were reliable with alpha = 0.78 and in the decreasing time condition alpha =0.82. The problems who invited percentage judgments above 100 were less consistent with decreasing distance alpha = 0.06 and for increasing time alpha = 0.65. We computed the mean judgments across items in each of the conditions omitting the decreasing distance items and called the variable means ***D***_***incr***_ (increasing distance), ***T***_***decr***_ (decreasing time) and ***T***_***incr***_ (increasing time). We then investigated co-variances between these variables and other items in the questionnaire. There were no significant correlations between the three variables across participants, but a marginally significant correlation between ***D***_***incr***_ and ***T***_***incr***_ (R =- 0.20, p=0.06).

### Additional questions

The mean response to the question “shortest distance from an infected person that would make you feel sufficiently safe” was 11.51 feet (SD=17.19), 3.51 meters. The Pearson correlations between the *shortest safe distance* and ***D***_***incr***_ and ***T***_***decr***_ were R = - 0.22 (p < 0.05) and R = 0.33 (p< 0.001) respectively. *Safe distance*, and ***T***_***incr***_ were unrelated. Generally speaking, this means that if a person wants a longer safety distance in a face to face conversation, she or he judges the effect of withdrawal on decrease of exposure as greater (relatively smaller percent judgments after change) than a person who needs only a shorter safety distance. Correspondingly, when a person wants a longer safety distance she or he judges the effect of shortening a conversation on exposure to be smaller (relatively greater percent judgments after change) than a person who states a shorter safety distance.

We asked: “assume a new Coronavirus epidemic occurred and no vaccine was available. Compared to the average person like yourself, how likely do think that it is that you would become sick? (Much less risk = 1, same risk as average person = 50, much greater risk =100)”. The median response was 50 meaning that overall there was no optimism bias (Svenson, 1981). We wanted to know if those judging themselves less likely to catch the disease were more optimistic than those who judged themselves as more likely to catch the disease were pessimistic. The mean response gives a hint about this question and it was 42.79 (SD = 24.66), which is significantly lower than 50, t (95)= -2.86, p >0.01 showing that the optimists were more optimistic than the pessimists were pessimistic. We asked if the respondent had been infected with a Corona virus to what extent would it depend on poor luck or poor behavior? The responses indicated influence from both factors with judgments around 50 (the midpoint of the VAS scale 1 - 100) with mean = 46.67 (SD = 28.33) and mean = 53.27 (SD=31.27) for the poor luck and poor behavior questions respectively. However, the difference is not statistically significant and these judgments had no correlations with optimism or pessimism.

As predicted, the question “How worried have you been over your own personal risk of becoming sick with COVID-19 during the pandemic?” correlated with “shortest acceptable distance”, R = 0. 23, p< 0.05, “avoiding public spaces”, R= 0.52, p<0.001 and “canceled or postponed meetings with friends” R = 0.49, p<0.001. These results corroborate the earlier findings (Bruine de Bruin & Bennett, 2020; Frounfelker et al.,2021; Shilo, Peleg & Nudelman, 2021; Vacondio et al., 2021). However, there were no significant correlations with this worry variable and ***D***_***incr***_ or ***T***_***decr***_. In summary, worry over catching COVID-19 was not directly related to judgments of the factual effects of changing distance and time of a conversation but to judgments of an acceptable distance to an infected person. We also asked about “general worry over things that may go wrong in life” but this unspecific worry did not correlate with protective behaviors.

## Study 2: Effects of distance and mask on judgments of exposure

In this study, we introduced problems with an infected person wearing a mask in a face to face conversation. The mask was a high quality mask that absorbed 90% of the exhaled particles.

### Method

#### Participants

In all, 150 participants were recruited by Prolific from a US adult English speaking population sample. They had not taken part in study1. After exclusion of 9 participants who gave judgments that implied that virus exposure was greater with than without a mask at the same distance, data from 141 participants remained for further analysis. There were 66 women, 74 men and 1 unspecified gender. The mean age was 32 years (SD= 13) with a range from 18 to 80 years. In the sample 1 had high school but no graduation, 15 were high school graduate, 42 had some college, 62 were college graduate and 21 had more than college graduate education.

### Procedure and Material

As in study 1, a Qulatrics questionnaire was distributed to the participants and a participant used on average 11 min to complete the task. We analyzed three questions about virus exposure comparing the protective effects of distance and face mask and two questions asking about the shortest distance from an infected person that the participant would accept as sufficiently safe. The three distance questions were presented in a random order that was unique for each participant and the other questions in appendix 1 were presented after them in a predetermined order. The introductory general instruction was as follows.

> *“As you probably know, the Corona virus (SARS-CoV2) spreads on small droplets and aerosols (very small airborne particles) in the air when a person infected with the virus breaths, coughs, sneezes or talks. Therefore, keeping a physical social distance and using a face mask both reduce virus exposure and the risk of the virus to spread from person to person. We will ask you to judge how different distances and/or a face mask can reduce your exposure to the virus when you do not wear a mask yourself in a face to face conversation with a person infected with COVID-19*.

All problems concerned conversations in an open space with no wind or draft and the instruction of the first problem included the following:

> *“If the infected person puts on a “90% face mask” (stopping 90% of exhaled viruses) you can move closer and yet keep the same safety standard because your exposure decreases. What is the closer distance to the infected person with a mask giving the same virus exposure as the one you accepted without a mask at 6 feet ?*
>
> *_____feet*_____*inches with a mask gives same level of exposure as at 6 feet without a 90% mask*.*”*

The second problem was the same but with 6 feet replaced by 11 feet. The third problem was introduced in this way.

> *“In another conversation the infected person without a mask moves away from you from 2 to 6 feet. What will the exposure be at 6 feet compared with the exposure at 2 feet? We set your Corona virus exposure at 2 feet to 100% virus exposure. When the other person moves away from you the initial 100 virus exposure decreases. But to what level does the exposure decrease?*
>
> *If you think that the exposure halves you write 50 and if it is one third 33 of the original exposure you write 33 percent, if it is one tenth 10 percent etc*
>
> *My total exposure after the moving away from 2 feet to 6 feet becomes*<u>____</u>*% of the 2 feet exposure”*

The face masks and distance problems were followed by a set of questions concerning the participants that were the same as in study 1 except the problems measuring cognitive ability. They were omitted because they did not co-vary with any of the other variables in study 1. In addition to the questions in study 1, we asked about the shortest inter-personal distance from an infected person that a participant could accept as sufficiently safe for a face to face 5 min conversation, with and without a mask on the infected person.

## Results

Typically, a high quality FFP2 mask stops 95% of the virus exposure if the mask is correctly fitted to the face (Howard et al., 2021) We decided to specify 90% efficiency in our problems to allow some mask misfit when we calculated predictions of correct solutions of the problems. A total of 29 participants had been diagnosed with Covid-19 but because they were such a small group they were not treated separately in the following analyses.

We will use meters as a measure of distance in Table 3 and in the following calculations. First, we denote the distance without a face mask *D*_*i*_ meters for the distance pair *i*. The distance with a mask that equals the protective effect of distance *D*_*i*_ is D_imask_. Then the mask reduction of exposure from 100% at D_i_ to 10% at D_imask;_ To repeat, without a mask the radiation is 100% at D_imask_ and the mask reduces exposure to 10%. We have to move away to Di without a mask to match that exposure. Then, according to Equation (1), with exposure decreasing with the square of the distance and (D_imask_/D_i_)^2^ = (10/100); D_imask_^2^ = 0.10 D_i_ ^2^; D_imask_ = 0.316 D_i_. Table 3 shows the predicted values for the first two problems. The linear estimates in the two first rows follow from (D_imask_/D_i_) = (10/100). The prediction of exposure when changing from 2 to 6 feet follows from n=2 in equation (1), (2/6)^2^ and for the linear prediction with the exponent 1.0.

**Table 3.** Judgments and objective estimates of the protective effects of interpersonal distance and mask against virus exposure.

|  | **Mean**<br>**(SD)** | **Median (m)**<br>**(Q<sub>25</sub> - Q<sub>75</sub>)** | **Predicted**<br>**VEM**<br>**n = 2.0** | **Predicted linear,**<br>**n = 1.0** | **Diff mean**<br>**judged -**<br>**predicted**<br>**VEM**<br>**n = 2.0** | **Diff median -**<br>**predicted**<br>**VEM**<br>**n = 2.0** |
| --- | --- | --- | --- | --- | --- | --- |
| **6 feet (1.83 m), closest with mask? N=140 (Q1)** | 1.24 m<br>(2.18) | 0.91 m<br>(0.61 - 1.63) | 0.58 m | 0.18 m | 0.66 m*** | 0.33 m |
| **11 feet (3.35 m), closest with mask? N=141 (Q2)** | 1.69 m<br>(1.97) | 1.64 m<br>(0.91 - 1.83) | 1.06 m | 0.17 m | 0.63 m*** | 0.58 m |
| **2 to 6 feet, (0.61 - 1.83 m) no mask percent remaining exposure? N=141 (Q7)** | 44.7% (37.8) | 33.0 %<br>(30 - 50) | 11.1% | 33.3% | 34.1%*** | 21.9 % |
| **Normal inter-personal distance without virus (G1)** | 3.35 feet<br>(1.91)<br>1.02 m | 3.00 feet<br>(2.00 - 4.00)<br>0.91 m |  |  |  |  |
| **Judged shortest safe distance without mask (G3)** | 11.6 feet<br>(10.40)<br>3.54 m | 10.0 feet<br>(6.00 - 12.00)<br>3.02 m |  |  |  |  |
| **Judged shortest safe distance with mask (G4)** | 7.10 feet<br>(4.42)<br>2.16 m | 6.00 feet<br>(4.00 - 9.00)<br>1.83 m |  |  |  |  |

For illustrative purposes, we tested the differences of the means from the predicted VEM 2.0 values for the three first rows in Table 3 in two tailed t tests. The test for the 6 feet distance problem resulted in t(139) = 7.84 p< 0.001, and the 11 feet problem t(140) = 6.05 p< 0.001. The median for the 2 to 6 feet approach, 33.0% was not too far from the median for the same problem in study 1 (30.0%) and the mean 44.7% was significantly different from the VEM 2.0 predicted value 11.1%, t(140) = 10.40 p< 0.001. Hence, the results show that the participants underestimated the effect of a face mask on reduction of virus exposure.

We also investigated the distribution of judgments to find out about the proportions of participants who made approximately correct judgments. For the 6 feet (1.83 m) problem we counted the number of participants who judged the distance to be smaller than 0.58 m In all, 35 participants (23%) gave distances that were equal or smaller than the predicted exposure. Hence, 77% underestimated the protective effect of a mask and selected a greater distance than required. For the 11 feet (3.35 m) problem with a cut point 1.06 m 45 (30%) of the participants gave distances that were equal to or smaller than the VEM 2.0 prediction, again indicating an underestimation of the protective effect of a mask. There were 11 participants (7%) who judged the exposure to be less than 13 % after a withdrawal from 2 to 6 feet and hence, 87% of the participants underestimated the protective effect of distance, which supports the results in study 1.

Now, we wanted to determine the perceived efficiency of a mask from the distance judgments in problems (1) and (2). The distance judgments given by the subjects include both the protective power of distance and mask, so we need to disentangle the distance effect from the mask effect. We will use problem (1) to illustrate how the effect of a mask can be estimated assuming no interaction effects between the distance and mask factors. We know from study 1 and the earlier study of Corona virus exposure (Svenson et al., 2020) that a linear VEM function predicts how judgments will describe the increase in exposure after coming closer to a person. This means that in problem (1) the judged approach from 1.83 m to 0.91 m gives a 1.83/0.91 = 2.01 increase in perceived exposure. Set the exposure at 1.83 m to 100% and then the exposure without a mask at 0.91 m will be 2.01 × 100%. The mask reduces the exposure 201% to 100%, that is (201 - 100)/201 = 50%. Then, the subjective efficiency of the mask gives a 50% reduction of virus exposure. This can be compared with the stated efficiency 90%. Hence, the participants underestimated the effect of a face mask. The corresponding analysis of problem (2) gives for an approach from 3.35 m to 1.64 m 3.35/1.64 = 2.04 increase in perceived exposure without a mask. Following the computation above, this problem indicates that the mask allows (204 - 100)/204 = 51%, of the exposure and the subjective efficiency is 49%, which again indicates underestimation of the actual efficiency.

The closest safe distances in Table 3 offers another way of estimating the subjective efficiency of a mask. Comparison of mean shortest distance without mask, 3.54 m and shortest distance with mask, 2.16 m indicates that the mask reduces the distance with 1.38 m which corresponds to the reduction of exposure. A subjective linear VEM decrease of exposure gives a remaining exposure 2.16/ 3.54 = 61% and the estimated efficiency of the mask is 39%.

The responses to the first two problems comparing situations with and without a mask correlated significantly R = 0.56, p < 0.001 and we decided to use the mean of the judgments in a search for covariance between the mask problems and other judgments.

However, there was only one marginally significant correlation between the means of the two mask problems and another variable, withdrawal from 2 feet to 6 feet, R = 0.16, p= 0.065.

The *withdrawal from 2 to 6 feet* responses correlated with *belief in following advice of distance and wearing a mask*, R(141) = - 0.19, p < 0.05. This means that a greater effect of withdrawal on exposure (smaller percentage of exposure) was positively associated with belief in the efficiency of following advice. The *belief in following advice* of *distance and wearing a mask* variable and *safety distance* correlated, R(141) = 0.24, p< 0.01. When a person indicated a greater safety distance, he or she had a stronger belief in following advice of distance and wearing a mask. As predicted from other research (Bruine de Bruin & Bennett, 2020; Frounfelker et al.,2021; Shilo, Peleg & Nudelman, 2021; Vacondio et al., 2021) *worry over becoming sick with Covid-19* correlated with *belief in following advice*, R (141) = 0.48, p <0.001.

In summary, a majority of the participants underestimated the effect of a face mask on reduction of virus exposure from 90% to about half of that percentage and underestimated the effect on virus exposure of moving away from an infected person.

## Discussion

As predicted, the studies showed that the effects on virus exposure of approaching and moving away from an infected person were underestimated. A linear instead of a curved relationship described the judgments, and hence explained the systematic underestimation bias. We used the exponent 2.0 in the VEM model to calculate the correct exposure values as a function of change of distance. This gives an underestimation in comparison with empirical studies of how quickly exposure changes with distance (Bjorn & Nielsen, 2002; Howard et l., 2021; Melikov, 2020). Therefore, when we reported that participants underestimated the change of exposure following a change of distance, this means that they were underestimating in relation to already conservative objective estimates of change of exposure. Hence, the reported underestimations may be even greater than what we have reported.

The normal inter individual distance in a face to face conversation, 1.91 m was increased to 3.54 m to make an average person feel safe when the other person was infected with a virus. We predicted that a person who selects a longer safety distance from an infected person should judge a change in interpersonal distance to have a smaller effect than a person who accepts a shorter safety distance. However, this prediction was not confirmed because safety distance was positively correlated with greater judgments of exposure change after change of distance. On average, those who were more sensitive to the effect of distance change selected longer safety distances. A participant who choose a relatively greater safety distance for her or himself judged the efficiency of official behavioral advice as relatively higher. When a participant wanted a longer safety distance, she or he judged the effect of shortening a conversation on exposure to be smaller than a person who indicted a shorter safety distance. These subjective relationships characterize the mental models that are elicited in response to the problems we have presented.

Our expectation that judgments of length of a conversation and exposure would be easier to judge correctly than distance was confirmed and the explanation that the linear relation relationship contributed to the correct judgments was confirmed. As predicted, worry of becoming sick with Covid-19 correlated positively with belief in following advice given by the authorities. The effect of a face mask on exposure reduction was underestimated in comparison with the mask performance given to the participants. In summary, the average participant revealed a mental model that included a linear relationship between distance and exposure and within this model (and objectively) the effect of wearing a face mask was underestimated. In general, people are not used to make the judgments that we have asked for, but our results indicate relationships in peoples’ mental models about airborne virus exposure that are relevant for behavior. We believe that this result is relevant for peoples’ actual protective behaviors when there is an ongoing epidemic disease or pandemic. For example, people including health workers (Atnafie et al., 2021) intuitively and subjectively may downplay the importance of keeping a sufficient distance and wearing a face mask. Then, their protective behavior depends on efficient facts and risk communications without an intuitive psychological foundation. Policy makers and politicians influenced by their intuitive understanding of the virus protective power of distancing and face masks, may hesitate to regulate their citizens’ behavior and not require distancing and face masks.

*

## Data Availability

All relevant data will be within the manuscript and its supporting Information files.

The author declares no potential competing interests. The study was approved by the ethics committee at Decision Research and the study was supported by the project Swedish Judgments at Decision Research.

## Acknowledgments

The author is grateful to Jeff Peterson for preparing the questionnaires and collecting the data and to Freja Isohani, Torun Lindholm and Henry Montgomery for valuable comments on an earlier version of the present contribution.

## Appendix 1

### Additional questions study 1

25. You have answered questions about inter-personal distance and virus exposure. What average distance do you keep to a person in a normal face to face conversation when no virus is around? I keep___meters
26. If you were infected with a new Corona virus, to what extent do you think that this could be caused by just you being unlucky or your own poor protective behavior? My poor luck_____(not at all = 0, only poor luck = 100) Poor protective behavior______(not at all = 0, only poor behavior = 100)
27. What is the shortest distance between you and a Corona virus infected person that would make you feel sufficiently safe to start a conversation of 3 minutes? _____meters would be sufficient for me to feel safe.
28. How worried have you been over your own personal risk of becoming sick with COVID-19 during the pandemic?_____ (0 = Not at all, 100 = Maximum)
29. In general, how worried are you over things that may go wrong in your life?______ (0 = Not at all, 100 = Maximum)
30. If you always follow the advice of keeping distance to other people. To what degree do you think that this behavior can protect you from being infected by a Corona virus assuming no mask ?_______ (0 = Not at all, 100 = Maximum)
31. How old are you?_____years
32. What gender ? Female______, Male_____, Other or no answer
33. Have you been diagnosed with COVID-19 by a test or by a doctor ? Yes____ No____
34. If you had the COVID - 19 infection and were sick, for how many days were you sick?_____ I have had no Covid infection____(mark applicable alternative)
35. Has anyone in your extended family been diagnosed with COVID - 19? (mark applicable alternative). Yes____ No____
36. If one person in your close family was sick, for how many days (the person who was most sick)?_____
37. Are you vaccinated against COVID - 19? Yes____I received the second dose_____(month) 2021 No or only one dose______(mark applicable alternative)
38. Assuming a new Corona virus epidemic without a vaccine, how likely do think that it is that you would be sick compared to the average person like yourself? My risk would be (scale 1 -100)_____ (Much less risk = 1, same risk as average person = 50, much greater risk =100)
39 If you should be infected with a new Corona virus without a vaccine, what is the risk that you would die from the infection?______ (No risk = 0, 100 = Certain that I would die) ***Which of the following have you done during the last seven days of the present Corona virus pandemic?***
40. Avoided public spaces, gatherings or crowds more often than before the pandemic? Never = 0, Always = 100 _______
41. Canceled or postponed meetings with friends more often than before the pandemic? ? Never = 0, Always = 100 _______
42. A bat and a ball cost $110 in total. The bat costs $100 more than the ball. How much does the ball cost? The cost of the ball in dollars: ______
43. If it takes 5 machines 5 minutes to make 5 widgets, how long would it take 100 machines to make 100 widgets? Number of minutes: ______
44. In a lake, there is a patch of lily pads. Every day, the patch doubles in size. If it takes 48 days for the patch to cover the entire lake, how long would it take for the patch to cover half of the lake? Number of days: ______
45. Please. indicate your highest level of education.

